# Association between stage-specific sleep bout durations and obstructive sleep apnea severity: A variable-domain functional regression approach

**DOI:** 10.64898/2026.07.14.26358060

**Authors:** Md Mushfiqur Rahman, Pratim Guha Niyogi

## Abstract

The apnea-hypopnea index (AHI), the conventional metric of obstructive sleep apnea (OSA) severity, is typically studied using scalar summaries of sleep architecture, such as the total time spent in each sleep stage. Although clinically interpretable, these summaries fail to capture the temporal organization of overnight sleep-stage sequences and may obscure stage-specific associations with OSA severity. Modeling the complete sleep-stage trajectory provides substantially richer temporal information; however, because total sleep duration varies across individuals, sleep-stage trajectories are observed over subject-specific domains, limiting the applicability of conventional functional regression methods that assume a common observation interval. We therefore applied Variable-Domain Functional Regression (VDFR) to overnight polysomnographic data from the APPLES study (*n* = 1, 103), treating the epoch-by-epoch sleep-stage sequence as a continuous, variable-length functional predictor of AHI. We compared three levels of sleep-stage granularity: five stages (Wakefulness, N1, N2, N3, REM), three stages (Wakefulness, Non-REM, REM), and binary staging (Wakefulness vs. Sleep). Functional sleep-stage terms were significant across all staging granularities and model structures (all p-values ≤ 0.001). Wake, N1, and N2 were positively associated with AHI, whereas N3 and REM were negatively associated, with REM exhibiting the strongest association. These effects were attenuated under coarser staging representations, highlighting the importance of preserving fine-grained sleep architecture. To our knowledge, this is the first application of VDFR to overnight polysomnographic data in OSA, showing that accommodating subject-specific sleep durations enables the identification of stage-specific temporal associations with AHI severity that are attenuated or obscured by coarser staging and conventional scalar analyses.

## Introduction

Obstructive sleep apnea (OSA) is a highly prevalent sleep disorder characterized by recurrent upper airway collapse during sleep, resulting in intermittent hypoxemia, sleep fragmentation, and repeated arousals [1]. Beyond its well-established cardiovascular and metabolic consequences [2–4], OSA has been associated with impairments in several neurocognitive domains, including attention, executive function, memory, and cognitive control, many of which depend on the integrity of the prefrontal cortex (PFC) [5, 6]. Despite substantial clinical interest, previous studies have reported inconsistent relationships between OSA severity and neurocognitive outcomes, suggesting that important aspects of the disease process remain insufficiently characterized [7, 8].

The Apnea Positive Pressure Long-term Efficacy Study (APPLES) is one of the largest multicenter investigations of OSA, collecting comprehensive overnight polysomnography (PSG) together with detailed neurocognitive assessments [9]. Analyses from APPLES and other cohorts have generally reported weak and inconsistent associations between the apnea-hypopnea index (AHI), the most widely used measure of OSA severity, and neurocognitive performance [6–8, 10]. While these studies have provided important insights, they have largely relied on scalar summaries of respiratory events and sleep architecture, potentially overlooking temporal features of sleep disruption that may be relevant to neurocognitive function.

Polysomnography provides an exceptionally rich temporal record of overnight sleep by continuously recording electroencephalography (EEG), electrooculography (EOG), electromyography (EMG), respiratory signals, and other physiological measurements [11, 12]. Sleep is scored on an epoch-by-epoch basis into Wake (W), three non-rapid eye movement (NREM) stages (N1, N2, and N3), and rapid eye movement (REM) sleep, producing a complete overnight sleep-stage sequence [11, 13]. Rather than representing isolated physiological states, these stages evolve through a highly organized temporal sequence, with repeated transitions throughout the night that are essential for normal sleep physiology. Growing evidence suggests that the neurophysiological functions of sleep depend not only on the total amount of time spent in each stage but also on the temporal organization and progression of these stages across the night [14–16].

In individuals with OSA, recurrent respiratory events disrupt the temporal organization of sleep by fragmenting sleep and selectively altering the occurrence of particular sleep stages, especially slow-wave sleep and REM sleep [14, 17, 18]. Nevertheless, conventional analyses typically summarize sleep architecture using aggregate measures, such as the total time or percentage of sleep spent in each stage. Although clinically interpretable, these summaries ignore the temporal organization of stage-specific sleep bout durations, reducing complex overnight sleep dynamics to a few scalar quantities. As a result, they cannot characterize how the timing and duration of sleep bouts within individual sleep stages are associated with OSA severity.

Functional data analysis (FDA) provides a natural framework for modeling physiological processes as continuous trajectories rather than scalar summaries [19–22]. By preserving the temporal structure of functional observations, FDA enables the investigation of how the evolution of a physiological process is associated with a clinical outcome. However, conventional functional regression methods assume that all trajectories are observed over a common fixed domain [19, 21, 23]. This assumption is violated in overnight PSG because total sleep duration varies substantially across individuals, resulting in sleep-stage trajectories with subject-specific domains.

To address this challenge, we employ Variable-Domain Functional Regression (VDFR), a statistical framework developed by [23] for scalar-on-function regression with functional predictors observed over subject-specific domains. Unlike conventional functional regression methods that assume all functional predictors are observed over a common domain, VDFR explicitly accommodates differences in domain length across individuals while preserving the temporal structure of the functional predictor. This makes it particularly well-suited for overnight PSG data, where total sleep duration naturally varies from one individual to another.

Although VDFR has been recognized as a promising framework for analyzing variable-length functional data and suggested as potentially useful for sleep research [23], to the best of our knowledge, it has not previously been applied to stage-specific sleep bout duration trajectories in OSA. Existing applications of functional data analysis in sleep research have primarily focused on continuous physiological signals, such as EEG spectral power [24] and actigraphy-derived activity profiles [22]. In contrast, studies investigating sleep architecture in OSA have largely relied on scalar summaries of PSG, including the total duration or proportion of time spent in individual sleep stages, rather than modeling the temporal organization of stage-specific sleep bout durations [25, 26].

In this study, we apply VDFR to overnight PSG data from the APPLES cohort to investigate the association between OSA severity, as measured by the apnea-hypopnea index (AHI), and the temporal organization of stage-specific sleep bout durations. By modeling sleep bout duration trajectories as variable-length functional predictors, rather than reducing sleep architecture to conventional scalar summaries, our approach preserves the temporal characteristics of sleep while accommodating differences in total sleep duration across individuals. This enables a more detailed characterization of how the temporal organization of sleep architecture is associated with OSA severity.

## Materials and methods

### Study population

Data for this analysis were drawn from the Apnea Positive Pressure Long-term Efficacy Study (APPLES), a randomized, double-blinded, sham-controlled, multicenter trial designed to evaluate the effect of continuous positive airway pressure (CPAP) on neurocognitive function in adults with obstructive sleep apnea [10]. A previously published paper [9] contains the detailed description of the APPLES protocol. De-identified baseline polysomnography (PSG) data were downloaded from the National Sleep Research Resource (NSRR; https://sleepdata.org), which provides public access to APPLES study data. The participants were recruited from five clinical sites across the United States. Clinically diagnosed OSA individuals with age ≥18 years as per American Academy of Sleep Medicine (AASM) were included in this study. The other inclusion criteria is, confirmed laboratory polysomnography (PSG) with an apnea-hypopnea index (AHI) ≥10 events/hour. In this current analysis, 1103 participants baseline epoch-level sleep stage data which was collected prior to randomization is used.

### Polysomnography

Overnight diagnostic polysomnography (PSG) was performed to continuously monitor physiological and electroencephalographic (EEG) signals using a standard comprehensive montage [10]. All recordings were electronically transmitted to a central Data Coordinating Center for standardized scoring.

Sleep architecture and wakefulness were initially scored using standard Rechtschaffen and Kales criteria [10, 27]. To construct the functional predictors for our variable-domain functional regression models, these historical structural classifications were mapped to the contemporary 5-stage categorizations: Wakefulness (W), Non-Rapid Eye Movement (NREM) stages N1, N2, and N3, and Rapid Eye Movement (REM) sleep [11]. For each participant, consecutive epochs belonging to the same sleep stage were grouped into individual sleep bouts. The duration of each bout was calculated as the total time spent in that uninterrupted stage before transitioning to a different stage. Separate stage-specific sleep bout duration functions were then constructed for Wake, N1, N2, N3, and REM. Figure 1 illustrates the construction of the functional predictors based on randomly selected subjects. Panel (A) displays the overnight sleep-stage hypnogram of a representative participant, while Panel (B) shows the corresponding stage-specific sleep bout durations, obtained by grouping consecutive epochs belonging to the same sleep stage. These stage-specific sleep bout duration functions were then used as the functional predictors in the subsequent variable-domain functional regression analyses.

**Fig 1.**
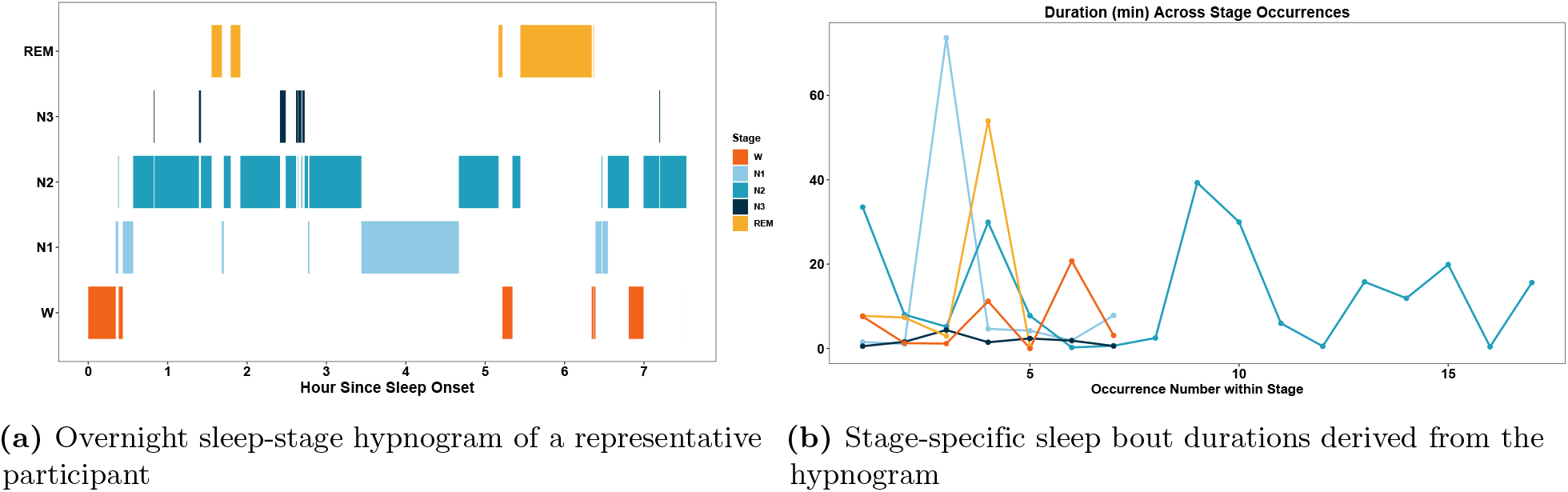
Construction of the stage-specific sleep bout duration functional predictors. (A) Overnight sleep-stage hypnogram of a representative participant obtained from polysomnography. (B) Stage-specific sleep bout durations were derived by grouping consecutive epochs belonging to the same sleep stage. These stage-specific sleep bout duration functions were subsequently used as the functional predictors in functional regression.

### Apnea-Hypopnea Index

The Apnea-Hypopnea Index (AHI) is defined as the total number of apneic and hypopneic events per hour of total sleep time, obtained from the baseline laboratory polysomnography [10, 28]. It is the most widely used clinical metric of OSA severity, with higher values indicating more frequent respiratory events during sleep [17]. An apnea was defined by a clear decrease of more than 90% from baseline in the amplitude of the nasal pressure signal lasting at least 10 seconds, while hypopneas were identified by a clear decrease of more than 50% from baseline in the nasal pressure signal associated with either an oxygen desaturation greater than 3% or an arousal [10]. OSA severity is conventionally classified according to AHI thresholds as mild (5 ≤ AHI < 15 events/hour), moderate (15≤ AHI < 30 events/hour), and severe (AHI ≥ 30 events/hour) [17, 28]. Larger AHI values indicate a greater frequency of respiratory events during sleep and, consequently, greater OSA severity. In the present analysis, the AHI was examined as a continuous outcome variable in a separate VDFR model, with the epoch-level sleep stage trajectory as the functional predictor, in order to investigate whether the temporal dynamics of sleep architecture carry information predictive of OSA severity beyond what is captured by conventional scalar summaries.

### Statistical analysis

To examine the association between OSA severity and stage-specific sleep bout duration functions, we employed Variable-Domain Functional Regression (VDFR), a scalar-on-function regression framework that accommodates functional predictors observed over subject-specific domains [23]. Stage-specific sleep bout duration functions were modeled as functional predictors because they preserve the temporal organization of sleep bouts throughout the night, rather than reducing them to aggregate scalar summaries. However, conventional functional regression methods assume that all functional predictors are observed over a common domain, an assumption that is violated in overnight PSG because total sleep duration naturally varies across individuals. A common strategy is to summarize sleep architecture using scalar measures, such as the total duration or percentage of time spent in each sleep stage, thereby discarding the temporal information contained in the sleep bout duration functions. Alternatively, forcing trajectories onto a common domain through temporal registration may distort their natural temporal characteristics. VDFR overcomes these limitations by directly modeling variable-length functional predictors while preserving their temporal structure.

The general VDFR model takes the form:

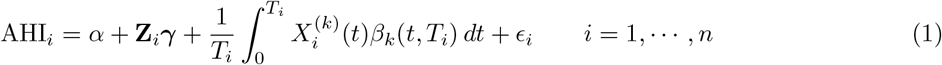

where for *i*-th individual, **Z**_*i*_ denotes the vector of non-functional covariates, 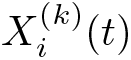 represents the stage-specific sleep bout duration function for the *k*-th sleep stage observed over the subject-specific domain [0, *T*_*i*_] for *i* = 1, *·, n* with sample size *n*. The term *β*_*k*_(*t, T*_*i*_) is the corresponding bivariate smooth coefficient function describing the association between the stage-specific functional predictor and AHI as a function of both the time within the sleep period and the participant’s total sleep duration. The scaling factor 1*/T*_*i*_ ensures that the functional effect is normalized with respect to the subject-specific domain, allowing coefficient estimates to remain comparable across participants with different sleep durations. The random error *ϵ*_*i*_ terms are assumed to be independent and normally distributed with mean zero and common variance *σ*^2^, where *σ*^2^ > 0.

An important property of the VDFR model is that it encompasses several widely used regression models as special cases. When all functional predictors are observed over a common domain, that is, *T*_*i*_ = *T* for all *i*, model (1) reduces to the classical scalar-on-function regression model [29, 30]. Furthermore, if the coefficient function is constant, i.e., *β*_*k*_(*t, T*_*i*_) = *β*_*k*_, for all *t*, then the model (1) reduces to a familiar linear regression model using the average value of the functional predictor.

In VDFR, the domain-standardized formulation was adopted for both computational and modeling purposes. Specifically, each subject’s functional predictor was linearly transformed to a common unit interval via the substitution *s* = *t/T*_*i*_, yielding standardized covariate functions 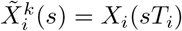 defined on [0, 1], where *s* represents the proportional position within the observation period. The model (1) then becomes: 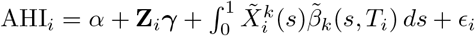where 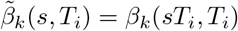 is the transformed bivariate coefficient function defined over the rectangular domain{ *s, T*_*i*_ : 0 ≤ *s* ≤ 1, 0 ≤ *T*_*i*_ ≤ max_*i*_ *T*_*i*_ .} This domain-standardized parameterization offers several practical advantages over the untransformed approach. First, it allows the use of an anisotropic tensor-product B-spline basis, which is well-suited to the rectangular domain of the standardized coefficient function and enables independent control of smoothness in the *s* and *T*_*i*_ directions. Second, it implicitly adjusts the degree of smoothness between adjacent time points relative to a subject’s total observation period, allowing greater flexibility for subjects with shorter follow-up. Third, it facilitates the implementation of parametric interaction structures, should a more parsimonious model be warranted.

The bivariate coefficient function 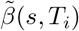 was estimated using a tensor-product B-spline basis with a second-order derivative penalty to ensure visual smoothness in both the *s* and *T*_*i*_ directions [31]. Model parameters were estimated via restricted maximum likelihood and linear mixed models [32]. All models were fit in R [33] using the `lf.vd()` function with the `transform = “standardized”`argument, called within the `pfr()` wrapper from the `refund` package [21, 23]. The estimated bivariate coefficient function 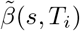 was examined to identify regions of the functional predictor domain that were significantly associated with the outcome, with pointwise confidence intervals obtained using standard sandwich estimators [21]. Regions where the pointwise 95% confidence interval excluded zero were considered statistically significant.

## Results

Table 1 presents the baseline characteristics of the 1,103 participants included in the analysis. The sample was predominantly male (65.55%) and largely White (76.16%), with a mean age of 51.56 years (SD = 12.21) and a mean of 15.50 years of education (SD = 2.59). Participants were, on average, obese, with a mean BMI of 32.21 kg/m^2^ (SD = 7.14) and mean weight of 212.05 lbs (SD = 49.87). The mean AHI was 40.09 events/hour (SD = 25.31), indicating a population with moderate to severe sleep-disordered breathing.

**Table 1.** Summary statistics of the study sample. For categorical variables, values are reported as the sample size with the corresponding proportion (percentage) in parentheses. For continuous variables, values are reported as the mean with the standard deviation in parentheses.

| Characteristic | n = 1,103 |
| --- | --- |
| Sex |  |
| Male | 723 (65.55%) |
| Female | 380 (34.45%) |
| Age | 51.56 (12.21) |
| Height | 68.10 (4.10) |
| Weight | 212.05 (49.87) |
| BMI | 32.21 (7.14) |
| Years of Education | 15.50 (2.59) |
| Marital Status |  |
| Single | 262 (23.75%) |
| Married | 639 (57.93%) |
| Other | 202 (18.31%) |
| Ethnicity |  |
| Asian | 61 (5.53%) |
| Black | 102 (9.25%) |
| White | 840 (76.16%) |
| Other | 100 (9.07%) |
| AHI | 40.09 (25.31) |

Table 2 presents the parametric coefficient estimates from the 5-stage VDFR model with AHI as the outcome. Corresponding results for the 3-stage and 2-stage models are provided in S1 Table. Across all three model structures, the parametric covariates demonstrated highly consistent patterns. Male sex was strongly and significantly associated with higher AHI across all sleep stages and all models (all *p <* 0.001), with coefficients ranging from approximately 6.67 to 8.44. BMI was similarly a robust positive predictor of AHI severity across all models and stages, with coefficients ranging from 1.27 to 1.41 (all *p <* 0.001). Age was also positively associated with AHI across most stages, though with smaller effect sizes (coefficients ranging from 0.14 to 0.21). In contrast, ethnicity and marital status were not significantly associated with AHI in any model, and years of education showed a modest but inconsistent negative association, reaching statistical significance only in the N2 stage of the 5-stage model (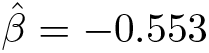, 95% CI: −1.071, −0.035) and the REM stage (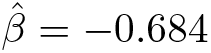, 95% CI: −1.219, −0.149).

**Table 2.**
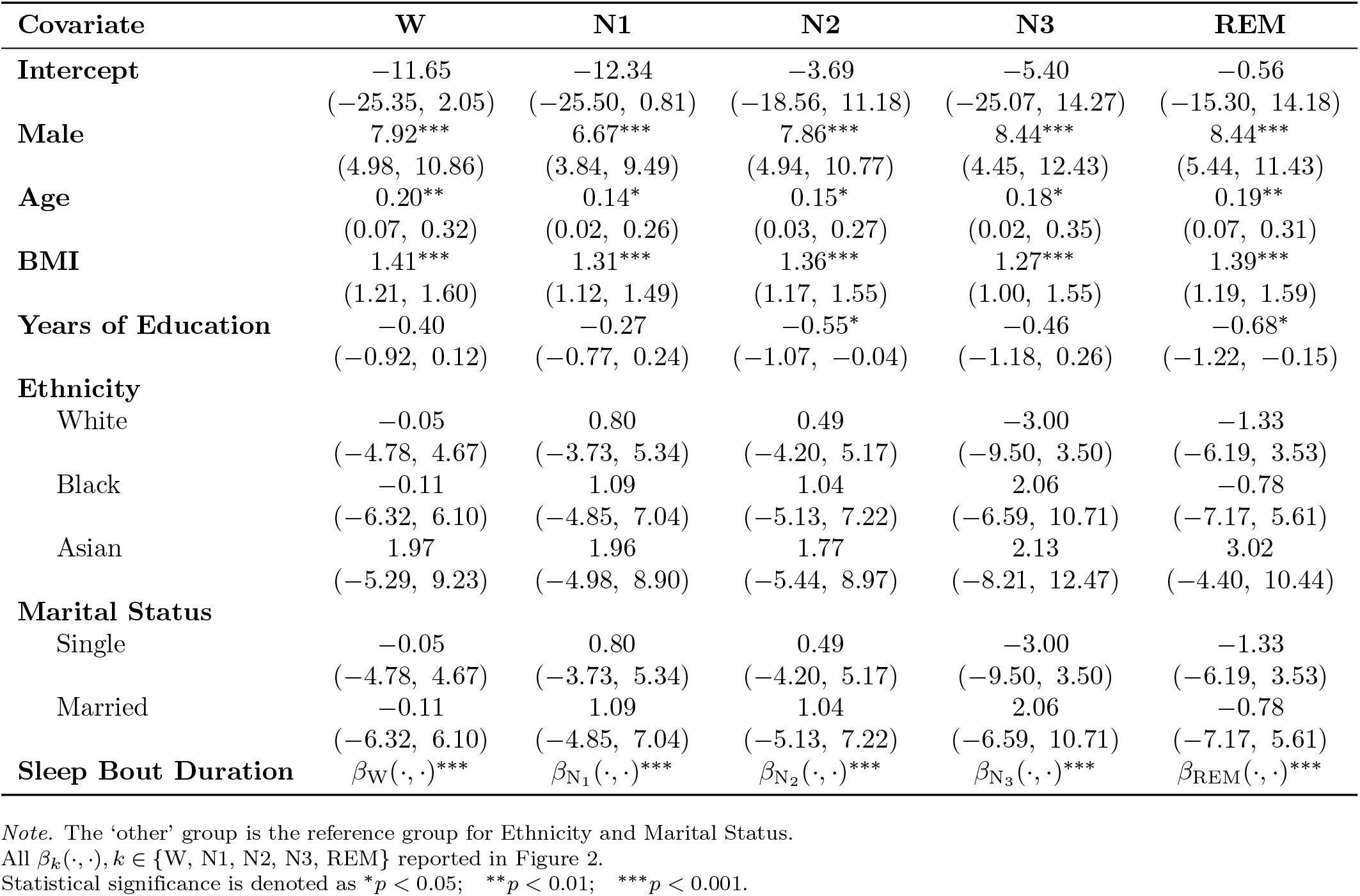
Regression coefficient estimates from the 5-stage variable-domain functional regression model with AHI as the continuous outcome. Each row corresponds to a separate model fitted using the stage-specific sleep bout duration function for Wake (W), Non-Rapid Eye Movement sleep stages (N1, N2, N3), or Rapid Eye Movement (REM) as the functional predictor. For the scalar covariates, regression coefficients are presented together with their corresponding 95% confidence intervals (in parentheses) and statistical significance.

Figure 2 presents the estimated bivariate coefficient surfaces 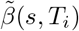 for the 5-stage model as heat maps. The horizontal axis (*t*) is the proportional position within the sleep period and the vertical axis (*T*_*i*_) denotes the participant-specific total sleep duration. Warmer colors (red/orange) represent areas of the sleep period where more time in a given stage is associated with higher AHI, and cooler colors (purple/blue) represent areas associated with lower AHI. To evaluate the robustness of the observed associations to the level of sleep-stage granularity, we additionally fitted analogous VDFR models using the conventional 3-stage (Wake, Non-REM, and REM) and 2-stage (Wake and Sleep) sleep classifications. The corresponding coefficient surfaces are presented in S1 Fig and S2 Fig, respectively.

**Fig 2.**
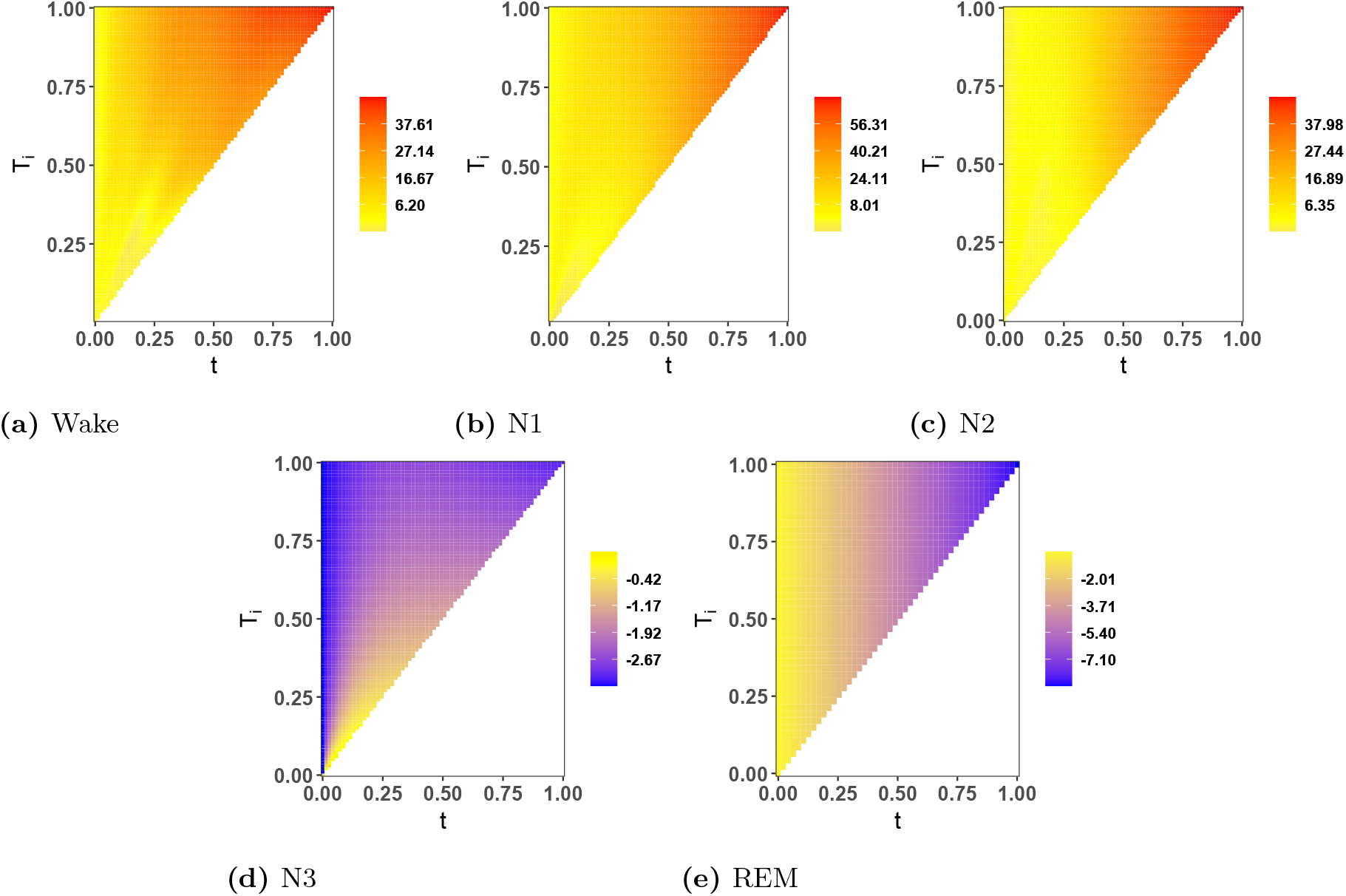
Functional regression coefficient estimates from the 5-stage variable-domain functional regression model with AHI. Each panel shows the estimated bivariate coefficient surface 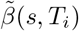 for a given sleep stage as a function of proportional time *t* ∈ [0, 1] (horizontal axis) and total sleep duration *T*_*i*_ (vertical axis). Warmer colors indicate stronger positive associations with AHI; cooler colors indicate negative associations. The white region (lower-right triangle) corresponds to values outside the variable domain *t* ≤ *T*_*i*_. Panels (A)–(E) correspond to Wake, N1, N2, N3, and Rapid Eye Movement (REM) sleep, respectively.

For Wake (W), the estimated coefficient surface was uniformly positive across most of the domain, with coefficient values ranging from 6.20 to 37.61. The magnitude of the coefficients increased toward the upper-right region of the surface, corresponding to later relative positions within the sleep period (*t* approaching 1.0) and longer total sleep durations (*T*_*i*_ near 1.0). These findings suggest that longer wake bout durations occurring later in the sleep period are more strongly associated with higher AHI, particularly among participants with longer sleep durations.

The 5-stage model exhibited consistently positive coefficient surfaces for both N1 and N2 sleep, with N1 creating the widest range of values (8.01 to 56.31) and the steepest gradient toward the upper-right region of the surface. This pattern suggests that later and longer sleep periods (N1 and N2 sleep) are the best predictors of higher AHI. In contrast to that, N3 (slow-wave sleep) and REM sleep possessed uniformly negative coefficient surfaces across the whole domain, N3 from -2.67 to -0.42 and REM from -7.10 to -2.01, which suggests that longer periods spent in these deeper stages, at any time during the sleep period and regardless of total sleep duration, are associated with lower AHI severity. The negative association was particularly strong for REM sleep, which had the most strongly negative and spatially consistent surface of all five stages.

These results collectively suggest that the temporal dynamics of sleep architecture are significantly and differentially associated with OSA severity. The directional consistency of the 5-stage coefficient surfaces, with lighter sleep stages (W, N1, N2) positively associated and deeper stages (N3, REM) negatively associated with AHI, implies that the temporal distribution of sleep stage composition throughout the night reflects meaningful variation in OSA severity. As shown in S1 Fig and S2 Fig, these opposing associations are reduced in the coarser 3-stage and 2-stage models, highlighting the value of finer staging granularity in capturing the full complexity of the sleep architecture and AHI relationship.

Overall, the comparison across staging granularities indicates that while the broad relationships between lighter and deeper sleep stages and AHI are robust, finer sleep-stage classifications provide substantially greater temporal resolution. Consequently, the 5-stage model better characterizes stage-specific associations with OSA severity than the conventional 3-stage and 2-stage classifications.

## Discussion

The opposite directional associations of AHI with the different sleep stages are physiologically coherent and consistent with the known tendency of OSA. The result shows that lighter sleep stages (Wakefulness, N1, and N2) were uniformly and positively associated with AHI, while deeper sleep stages (REM and N3) were negatively associated with AHI. This is in line with the pattern of selective fragmentation and suppression of slow wave and REM sleep in OSA, and the increased representation of lighter NREM stages and wakefulness throughout the night.

Moreover, such associations were most prominent at later proportional time points in the sleep period and in subjects with longer total sleep times, a characteristic that would be completely missed by conventional scalar approaches such as the AHI or stage-specific percentage summaries. The strong and spatially uniform negative relationship of REM sleep with AHI across the entire coefficient surface is consistent with the well-established susceptibility of REM sleep to respiratory disruption in OSA and with the more general role of REM sleep in cognitive restoration and emotional regulation [14–16]. Similarly, the positive associations of N1 and N2 with AHI severity, where the strongest association is in the later part of sleep, are in alignment with the clinical observation that lighter NREM sleep occurs most frequently in the second half of the night in OSA, when REM pressure accumulates, and respiratory drive is gradually diminished. [17, 18].

A secondary, but important finding is with respect to the role of staging granularity. The coefficient surfaces of the 5-stage model were consistently more detailed and informative than the 3-stage or binary models. In particular, combining N1, N2, and N3 into a single NREM category in the 3-stage model obscured inverse directional relationships of lighter versus deeper NREM sleep with AHI severity, thus reducing the sensitivity of the model to clinically relevant shifts in sleep architecture. Similarly, although the binary model maintained statistically significant functional associations, it necessarily lost the stage-specific resolution required to determine which components of sleep disruption are most strongly associated with OSA severity. These results suggest that the full granularity of sleep stages should be preserved to capture the full relationship between sleep architecture dynamics and OSA. More generally, they are a warning against the use of coarse staging schemes as a routine part of functional analyses of PSG data, as loss of information at the staging level will propagate directly to attenuation of the estimated functional associations. VDFR has some methodological advantages over the traditional approaches to analyze PSG data in OSA research. By allowing for the presence of subject-specific variation in the total sleep duration, a salient feature of OSA populations, without the need for artificial domain registration, and by estimating smooth bivariate coefficient surfaces that characterize the effects of both the timing and the total duration of sleep, VDFR offers a principled and flexible framework for leveraging the full temporal richness of overnight In PSG recordings [23], the effects of parametric covariates were highly consistent across all model structures, with male sex and BMI being the dominant predictors of AHI severity [17], providing further evidence of the robustness of the approach and corroborating established risk factor profiles for OSA in a large community-based cohort [3, 4].

Some limitations of the present study are there. First, as the analysis was cross-sectional, we cannot infer causality for the direction of the relationships we observed between the dynamics of sleep architecture and OSA severity. Second, the APPLES cohort was relatively highly educated and largely white, which may limit the generalizability of these findings to more diverse populations. Future work should explore the longitudinal stability of these functional associations, explore whether VDFR-derived coefficient surfaces can be used as clinically useful biomarkers of OSA severity, and extend the framework by incorporating additional physiological signals available from the PSG recording, such as oxygen saturation trajectories and EEG spectral features.

## Conclusion

VDFR regarded the sequence of overnight sleep stages as a continuous functional object instead of a scalar summary of sleep and revealed temporally structured and stage-specific associations between the dynamics of sleep architecture and OSA severity that are invisible to conventional approaches. Lighter stages (Wakefulness, N1, and N2) were found to be consistently positively associated with AHI, whereas deep stages (N3 and RWM) were negatively associated. Also, the effect was largest for individuals who slept longer and in the later part of sleep, in the case of deep stages. These method of analyzing sleep stage duration data and the results highlights the value of preserving the fine-grained, moment-to-moment details of sleep study data, offering a promising new way to better understand the complex links between sleep patterns and the real-world health impacts of OSA.

## Data Availability

All data produced in the present study are available upon reasonable request to the authors. The data used in this article is publicly available at https://sleepdata.org.

## Supporting information

**S1 Fig. Functional coefficient estimates from the 3-stage variable-domain functional regression model with AHI**. Each panel shows the estimated bivariate coefficient surface 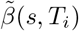 as a function of the proportional time *t* and total sleep duration *T*_*i*_. The combined NREM stage (N) showed a positive coefficient surface (values: 5.26 to 27.07) with a pattern somewhat intermediate between the positive N1/N2 and negative N3 surfaces from the 5-stage model, reflecting the averaging effect of lumping all three NREM substages into one category. REM surface was identical to that of the 5-stage model (values: − 7.10 to −2.01). This comparison highlights a key limitation of the 3-stage granularity, namely, that by collapsing N1, N2, and N3 into a single NREM category, the opposing directional associations of lighter versus deeper NREM sleep with AHI severity were obscured, which may have decreased the sensitivity of the model to clinically meaningful differences in sleep architecture.

**S2 Fig. Functional coefficient estimates from the 2-stage variable-domain functional regression model with AHI**. The estimated bivariate coefficient surface 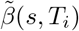 is plotted as a function of proportional time *t* (horizontal axis) and total sleep duration *T*_*i*_ (vertical axis) in each panel. The Non-Wake (NW) stage in the 2-stage model showed a similar pattern to the Wakefulness surface in the 5-stage model (values: 4.85 to 22.13), suggesting a positive association between overall sleep time, regardless of specific stage, and AHI. Although the 2-stage model preserved statistically significant functional associations, it necessarily lost stage-specific resolution that would help to identify which components of sleep disruption are most strongly associated with OSA severity.

**S1 Table. Estimates (95% CI) from variable-domain functional regression models with AHI across the 3-stage, and 2-stage model**. Full parametric coefficient estimates and 95% confidence intervals from the 3-stage, and 2-stage VDFR models with AHI as the outcome. Parametric covariate effects were highly consistent across all three model structures.

## Notes

### Competing Interest Statement

The authors have declared no competing interest.

### Author Declarations

De-identified data were downloaded from the National Sleep Research Resource (NSRR; https://sleepdata.org), which provides public access to APPLES study data.

## References

1. Young T, Palta M, Dempsey J, Skatrud J, Weber S, Badr S. The Occurrence of Sleep-Disordered Breathing among Middle-Aged Adults. New England Journal of Medicine. 1993;328(17):1230–5.

2. Peker Y, Hedner J, Norum J, Kraiczi H, Carlson J. Increased Incidence of Cardiovascular Disease in Middle-Aged Men with Obstructive Sleep Apnea: A 7-Year Follow-Up. American Journal of Respiratory and Critical Care Medicine. 2002;166(2):159–65.

3. Punjabi NM, Caffo BS, Goodwin JL, et al. Sleep-Disordered Breathing and Mortality: A Prospective Cohort Study. PLoS Medicine. 2009;6(8):e1000132.

4. Redline S, Yenokyan G, Gottlieb DJ, et al. Obstructive Sleep Apnea Hypopnea and Incident Stroke: The Sleep Heart Health Study. American Journal of Respiratory and Critical Care Medicine. 2010;182(2):269–77.

5. Beebe DW, Gozal D. Obstructive Sleep Apnea and the Prefrontal Cortex: Towards a Comprehensive Model Linking Nocturnal Upper Airway Obstruction to Daytime Cognitive and Behavioral Deficits. Journal of Sleep Research. 2002;11(1):1–16.

6. Engleman HM, Douglas NJ. Sleep· 4: Sleepiness, Cognitive Function, and Quality of Life in Obstructive Sleep Apnoea/Hypopnoea Syndrome. Thorax. 2004;59(7):618–22.

7. Beebe DW, Groesz L, Wells C, Nichols A, McGee K. The Neuropsychological Effects of Obstructive Sleep Apnea: A Meta-Analysis of Norm-Referenced and Case-Controlled Data. Sleep. 2003;26(3):298–307.

8. Weaver TE. Outcome Measurement in Sleep Medicine Practice and Research. Part 2: Assessment of Neurobehavioral Performance and Mood. Sleep Medicine Reviews. 2001;5(3):223–36.

9. Kushida CA, Nichols DA, Quan SF, Goodwin JL, White DP, Gottlieb DJ, et al. The Apnea Positive Pressure Long-term Efficacy Study (APPLES): Rationale, Design, Methods, and Procedures. Journal of Clinical Sleep Medicine. 2006;2(03):288–300.

10. Quan SF, Chan CS, Dement WC, Gevins A, Goodwin JL, Gottlieb DJ, et al. The Association between Obstructive Sleep Apnea and Neurocognitive Performance—The Apnea Positive Pressure Long-term Efficacy Study (APPLES). Sleep. 2011;34(3):303–14.

11. Berry RB. The AASM Manual for the Scoring of Sleep and Associated Events: Rules, Terminology and Technical Specifications. Darien, IL: American Academy of Sleep Medicine; 2014.

12. Iber C, Ancoli-Israel S, Chesson AL, Quan SF. The AASM Manual for the Scoring of Sleep and Associated Events: Rules, Terminology and Technical Specifications. 1st ed. Westchester, IL: American Academy of Sleep Medicine; 2007.

13. Carskadon MA, Dement WC, et al. Normal Human Sleep: An Overview. Principles and Practice of Sleep Medicine. 2005;4(1):13–23.

14. Diekelmann S, Born J. The Memory Function of Sleep. Nature Reviews Neuroscience. 2010;11(2):114–26.

15. Walker MP. The Role of Sleep in Cognition and Emotion. Annals of the New York Academy of Sciences. 2009;1156(1):168–97.

16. Rasch B, Born J. About Sleep’s Role in Memory. Physiological Reviews. 2013;93(2):681–766.

17. Punjabi NM. The Epidemiology of Adult Obstructive Sleep Apnea. Proceedings of the American Thoracic Society. 2008;5(2):136–43.

18. Born J, Rasch B, Gais S. Sleep to Remember. The Neuroscientist. 2006;12(5):410–24.

19. Ramsay JO, Silverman BW. Functional Data Analysis. New York: Springer; 2005.

20. Morris JS. Functional Regression. Annual Review of Statistics and Its Application. 2015;2:321–59.

21. Goldsmith J, Bobb J, Crainiceanu CM, Caffo B, Reich D. Penalized Functional Regression. Journal of Computational and Graphical Statistics. 2011;20(4):830–51.

22. Di CZ, Crainiceanu CM, Caffo BS, Punjabi NM. Multilevel Functional Principal Component Analysis. The Annals of Applied Statistics. 2009;3(1):458–88.

23. Gellar JE, Colantuoni E, Needham DM, Crainiceanu CM. Variable-Domain Functional Regression for Modeling ICU Data. Journal of the American Statistical Association. 2014;109(508):1425–39.

24. Crainiceanu CM, Caffo BS, D. CZ, Punjabi NM. Nonparametric Signal Extraction and Measurement Error in the Analysis of Electroencephalographic Activity During Sleep. Journal of the American Statistical Association. 2009;104(486):541–55.

25. Blackwell T, Yaffe K, Laffan A, Ancoli-Israel S, Redline S, Ensrud KE, et al. Associations of Objectively and Subjectively Measured Sleep Quality with Subsequent Cognitive Decline in Older Community-Dwelling Men: The MrOS Sleep Study. Sleep. 2011;34(10):1347–56.

26. Mander BA, Winer JR, Walker MP. Sleep and Human Aging. Neuron. 2017;94(1):19–36.

27. Rechtschaffen A. A Manual of Standardized Terminology, Techniques and Scoring System for Sleep Stages of Human Subjects. Washington, DC: US Government Printing Office, National Institute of Health; 1968.

28. American Academy of Sleep Medicine Task Force. Sleep-Related Breathing Disorders in Adults: Recommendations for Syndrome Definition and Measurement Techniques in Clinical Research. Sleep. 1999;22(5):667–89.

29. Reiss PT, Goldsmith J, Shang HL, Ogden RT. Methods for scalar-on-function regression. International Statistical Review. 2017;85(2):228–49.

30. Guha Niyogi P, Dhar SS. Identifying arbitrary transformation between the slopes in scalar-on-function regression. Electronic Journal of Statistics. 2026;20(1):2211–53.doi:10.1214/26-EJS2535.

31. Wood SN. Thin Plate Regression Splines. Journal of the Royal Statistical Society: Series B (Statistical Methodology). 2003;65(1):95–114.

32. Ruppert D, Wand MP, Carroll RJ. Semiparametric Regression. 12. Cambridge University Press; 2003.

33. R Core Team. R: A Language and Environment for Statistical Computing. Vienna, Austria; 2025. Available from: https://www.R-project.org/.

