## Supplementary Figure 1 for "Association between stage-specific sleep bout durations and obstructive sleep apnea severity: A variable-domain functional regression approach"

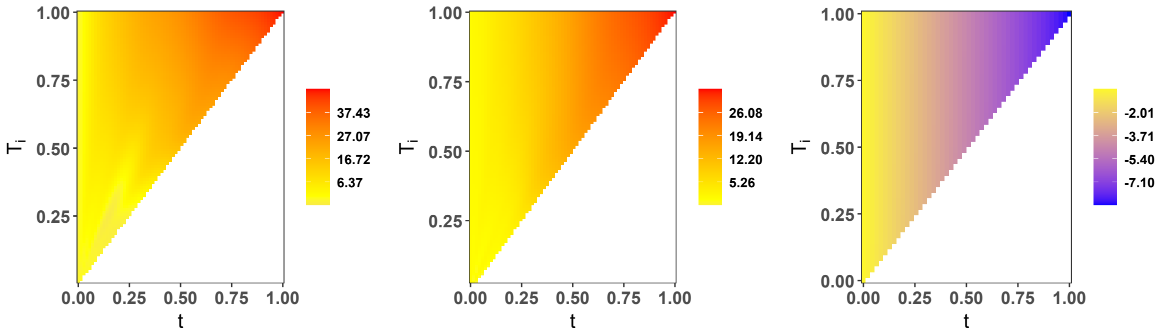


(a) Wake (b) Non-REM (c) REM

**S1 Fig.** Functional regression coefficient estimates from the 3-stage variable-domain functional regression model with AHI. Each panel shows the estimated bivariate coefficient surface $\tilde{\beta}(s, T_i)$ for a given sleep stage as a function of proportional time $t \in [0, 1]$ (horizontal axis) and total sleep duration $T_i$ (vertical axis). Warmer colors indicate stronger positive associations with AHI; cooler colors indicate negative associations. The white region (lower-right triangle) corresponds to values outside the variable domain $t \leq T_i$. Panels (A)–(C) correspond to Wake, Non-Rapid Eye Movement (NREM) and Rapid Eye Movement (REM) sleep, respectively.
