## Supplementary Table 1 for "Association between stage-specific sleep bout durations and obstructive sleep apnea severity: A variable-domain functional regression approach"

**S1 Table.** Regression coefficient estimates from the 5-stage variable-domain functional regression model with AHI as the continuous outcome. Each row corresponds to a separate model fitted using the stage-specific sleep bout duration function as the functional predictor under the corresponding staging granularity: Wake (W), Non-Rapid Eye Movement (NREM) sleep, and REM sleep (3-stage); or Wake (W) and Non-Wake (NW) (2-stage). For the scalar covariates, regression coefficients are presented together with their corresponding 95% confidence intervals (in parentheses) and statistical significance.

| | **3-Stage Model** | | | **2-Stage Model** | |
|---|---|---|---|---|---|
| **Covariate** | **W** | **N** | **REM** | **W** | **NW** |
| **Intercept** | $-10.59$ | $-25.62^{**}$ | $-0.56$ | $-11.65$ | $-16.96^{*}$ |
| | $(-24.36, 3.17)$ | $(-41.31, -9.94)$ | $(-15.30, 14.18)$ | $(-25.35, 2.05)$ | $(-33.26, -0.67)$ |
| **Male** | $7.64^{***}$ | $7.38^{***}$ | $8.44^{***}$ | $7.92^{***}$ | $7.54^{***}$ |
| | $(4.69, 10.59)$ | $(4.48, 10.29)$ | $(5.44, 11.43)$ | $(4.98, 10.86)$ | $(4.61, 10.47)$ |
| **Age** | $0.21^{***}$ | $0.19^{**}$ | $0.19^{**}$ | $0.20^{**}$ | $0.16^{*}$ |
| | $(0.09, 0.34)$ | $(0.07, 0.31)$ | $(0.07, 0.31)$ | $(0.07, 0.32)$ | $(0.03, 0.28)$ |
| **BMI** | $1.38^{***}$ | $1.32^{***}$ | $1.39^{***}$ | $1.41^{***}$ | $1.37^{***}$ |
| | $(1.18, 1.57)$ | $(1.13, 1.52)$ | $(1.19, 1.59)$ | $(1.21, 1.60)$ | $(1.18, 1.57)$ |
| **Years of Education** | $-0.41$ | $-0.37$ | $-0.68^{*}$ | $-0.40$ | $-0.52$ |
| | $(-0.93, 0.12)$ | $(-0.88, 0.15)$ | $(-1.22, -0.15)$ | $(-0.92, 0.12)$ | $(-1.04, 0.00)$ |
| **Ethnicity** | | | | | |
| White | $-0.10$ | $0.87$ | $-1.33$ | $-0.05$ | $1.76$ |
| | $(-4.81, 4.62)$ | $(-3.78, 5.52)$ | $(-6.19, 3.53)$ | $(-4.78, 4.67)$ | $(-2.97, 6.50)$ |
| Black | $-0.38$ | $0.80$ | $-0.78$ | $-0.11$ | $2.15$ |
| | $(-6.62, 5.86)$ | $(-5.37, 6.97)$ | $(-7.17, 5.61)$ | $(-6.32, 6.10)$ | $(-4.09, 8.39)$ |
| Asian | $2.06$ | $3.37$ | $3.02$ | $1.97$ | $3.78$ |
| | $(-5.18, 9.30)$ | $(-3.75, 10.48)$ | $(-4.40, 10.44)$ | $(-5.29, 9.23)$ | $(-3.42, 10.98)$ |
| **Marital Status** | | | | | |
| Single | $-0.10$ | $0.87$ | $-1.33$ | $-0.05$ | $1.76$ |
| | $(-4.81, 4.62)$ | $(-3.78, 5.52)$ | $(-6.19, 3.53)$ | $(-4.78, 4.67)$ | $(-2.97, 6.50)$ |
| Married | $-0.38$ | $0.80$ | $-0.78$ | $-0.11$ | $2.15$ |
| | $(-6.62, 5.86)$ | $(-5.37, 6.97)$ | $(-7.17, 5.61)$ | $(-6.32, 6.10)$ | $(-4.09, 8.39)$ |
| **Sleep Bout Duration** | $\beta_{\mathrm{W}}(\cdot,\cdot)^{***}$ | $\beta_{\mathrm{N}}(\cdot,\cdot)^{***}$ | $\beta_{\mathrm{REM}}(\cdot,\cdot)^{***}$ | $\beta_{\mathrm{W}}(\cdot,\cdot)^{***}$ | $\beta_{\mathrm{NW}}(\cdot,\cdot)^{***}$ |

*Note.* The 'other' group is the reference group for Ethnicity and Marital Status.
All $\beta_k(\cdot,\cdot), k \in \{\mathrm{W, N, REM}\}$ or $k \in \{\mathrm{W, NW}\}$ based on the granularity reported in Figures S1 and S2, receptively.
Statistical significance is denoted as $^{*}p < 0.05$; $^{**}p < 0.01$; $^{***}p < 0.001$.
