## Supplementary Figure 2 for "Association between stage-specific sleep bout durations and obstructive sleep apnea severity: A variable-domain functional regression approach"

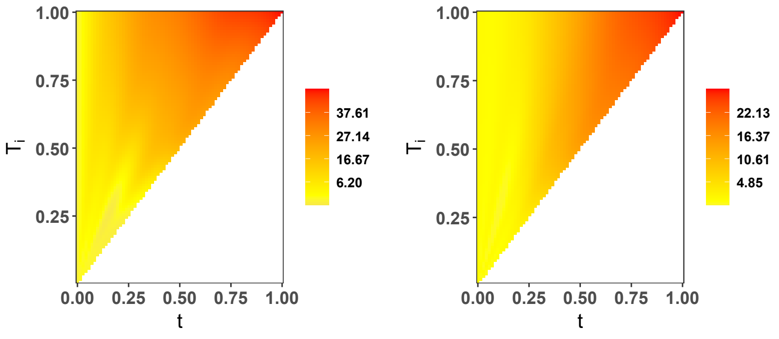


(a) Wakefulness (b) Non-wakefulness

**S2 Fig.** Functional regression coefficient estimates from the 2-stage variable-domain functional regression model with AHI. Each panel shows the estimated bivariate coefficient surface $\tilde{\beta}(s, T_i)$ for a given sleep stage as a function of proportional time $t \in [0, 1]$ (horizontal axis) and total sleep duration $T_i$ (vertical axis). Warmer colors indicate stronger positive associations with AHI; cooler colors indicate negative associations. The white region (lower-right triangle) corresponds to values outside the variable domain $t \leq T_i$. Panels (A)–(B) correspond to Wakefulness and non-wakefulness, respectively.
